## Supplementary Information for "Cutaneous and acral melanoma cross-OMICs reveals prognostic cancer drivers associated with pathobiology and ultraviolet exposure"

### Supplementary Figures

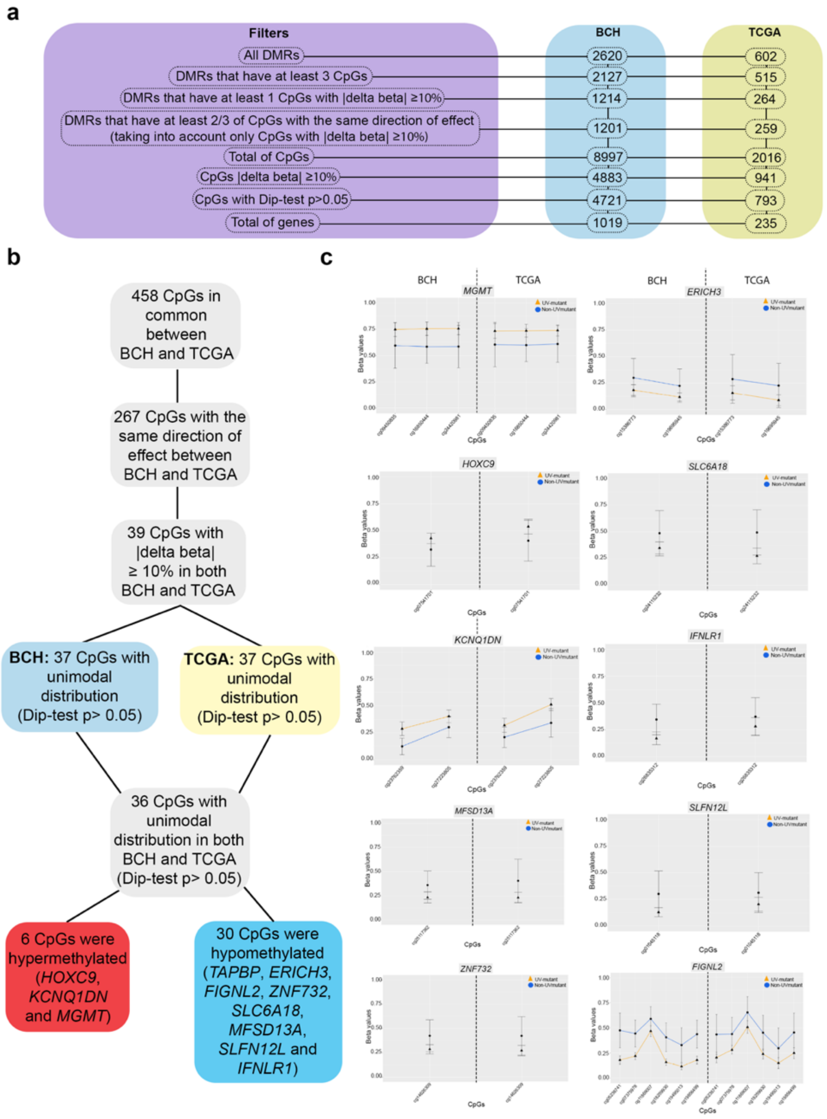


**Supplementary Figure 1:** Prioritization of UV methylation markers. a) Filtration steps applied to DMRs derived from the crude model in order to select the top CpGs and genes that are differentially methylated between UV-mutant *versus* non UV-mutant in BCH and TCGA. Significance was assessed using linear robust regression with FDR < 0.05. b) Prioritization criteria of UV-associated CpGs that are differentially methylated in both BCH and TCGA. c) DNA methylation levels of the 9 genes in common between BCH and TCGA showing differential methylation between UV-mutant (n= 44 and 47 in BCH and TCGA, respectively) and non UV-mutant patients (n= 44 and 47 in BCH and TCGA, respectively). Data were expressed as the average values of each group (UV-mutant and non UV-mutant) for each single CpG with error bars indicating the 95% confidence interval.

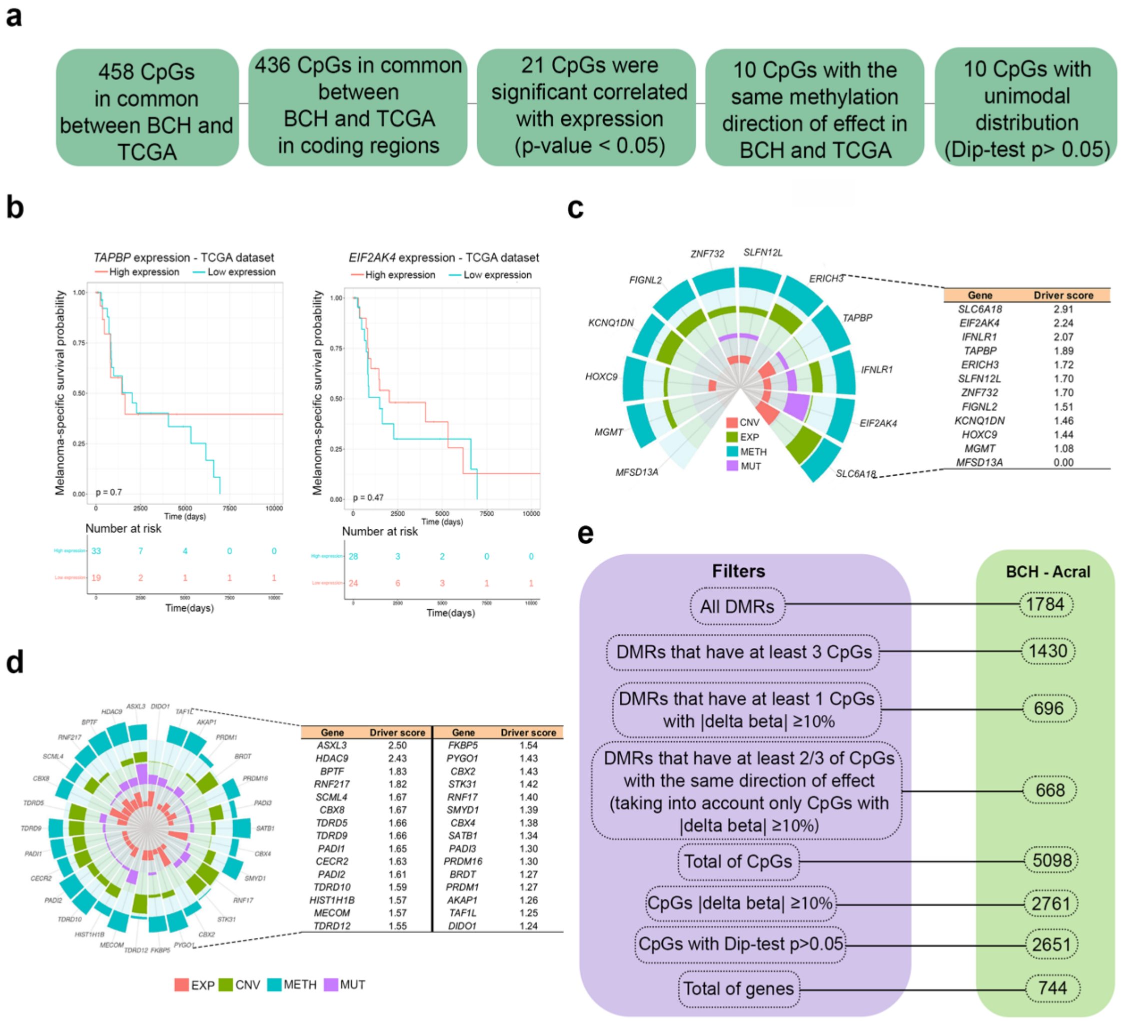


**Supplementary Figure 2:** Association of DNA methylation with transcription, transcription-mediated patient survival, cancer driver potential and pathobiology. a) Prioritization criteria of CpGs with significant expression quantitative trait methylation (eQTMs) ($p < 0.05$) in both BCH and TCGA. P-value was delivered from two-sided Pearson correlation and Dip-tests. b) Kaplan-Meier survival of melanoma patients in relation to expression levels of *TAPBP* and *EIF2AK4* measured in the target tumors derived from TCGA. Patients were categorized into low- and high-expression groups relative to the mean value of expression across profiled samples for a given gene. P values were derived by log-rank test. c) and d) Multi-omics data integration, encompassing copy number variation (CNV), expression (EXP), methylation (METH) and mutation (MUT), was performed in order to decipher the melanoma driver potential of the 12 prioritized genes (c) and of positive control genes previously identified in a recent study based on the ConsensusDriver method[1] (d). e) Filtration steps applied to DMRs derived from the crude model in order to select the top CpGs and genes that are differentially methylated between cutaneous and acral melanoma patients not harboring the UV mutational signature.

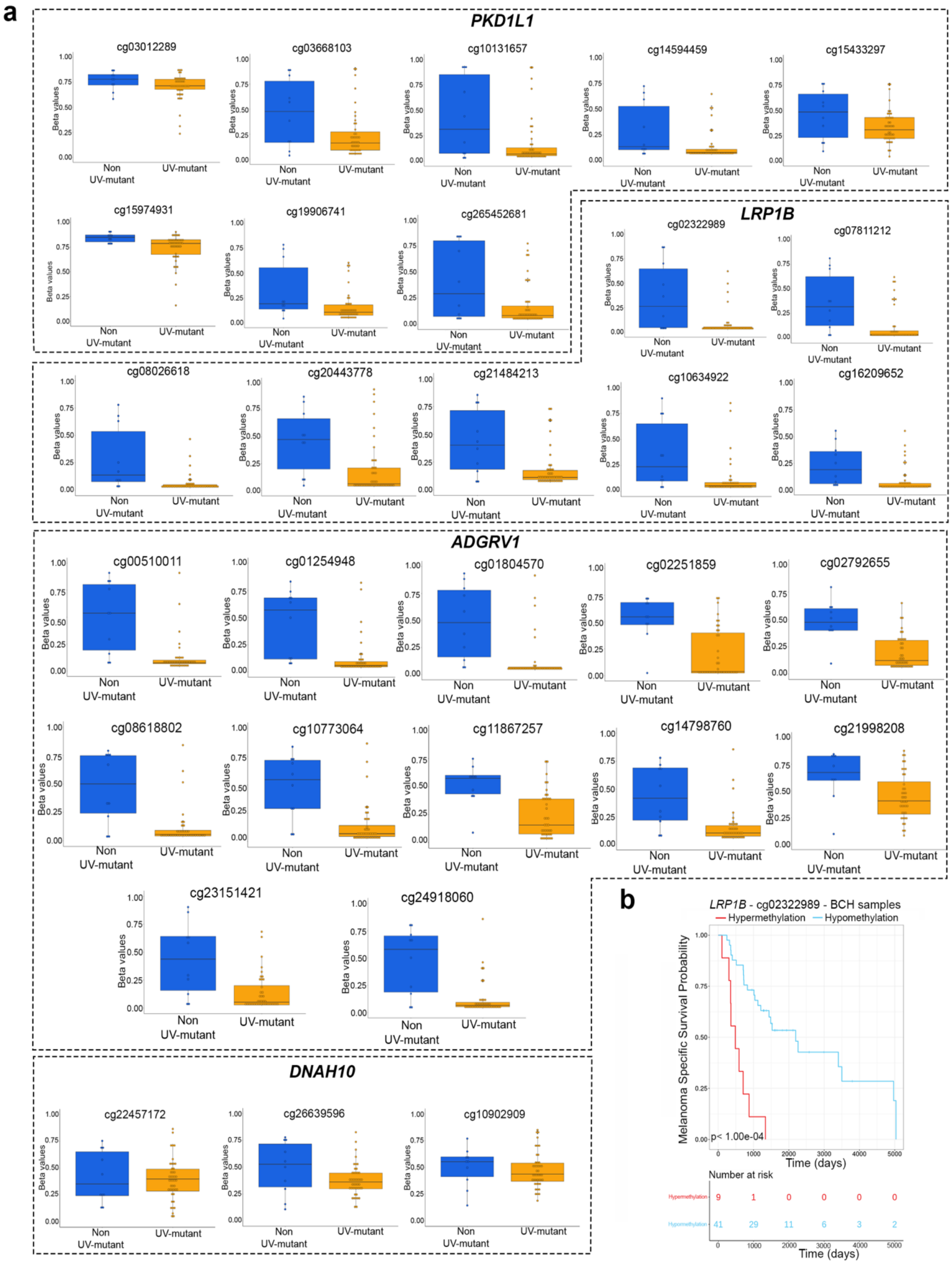


**Supplementary Figure 3**: DNA methylation alterations and clinical relevance of genes reported in the literature to be frequently mutated in UV-mutant *versus* non UV-mutant cutaneous melanoma patients[2]. a) *LRP1B*, *PKHD1L1*, *ADGRV1* and *DNAH10* were differentially methylated in UV-mutant (n= 44) relative to non UV-mutant (n= 10) patients in the BCH-cutaneous cohort. Box center lines, bound of the box, and whiskers indicate medians, first and third quantiles, and minimum and maximum values within 1.5xIQR (interquartile range) of the box limits, respectively. Each data point in the box plot represents the samples. b) Kaplan-Meier survival of cutaneous melanoma patients in relation to methylation levels of cg02322989 (*LRP1B*) measured in the target tumors derived from BCH. Patients were categorized into low- and high-methylation groups depending on whether the methylation value of a given CpG is lower or higher, respectively, than the mean methylation across the samples profiled for that CpG. The P-value was derived by log-rank test.

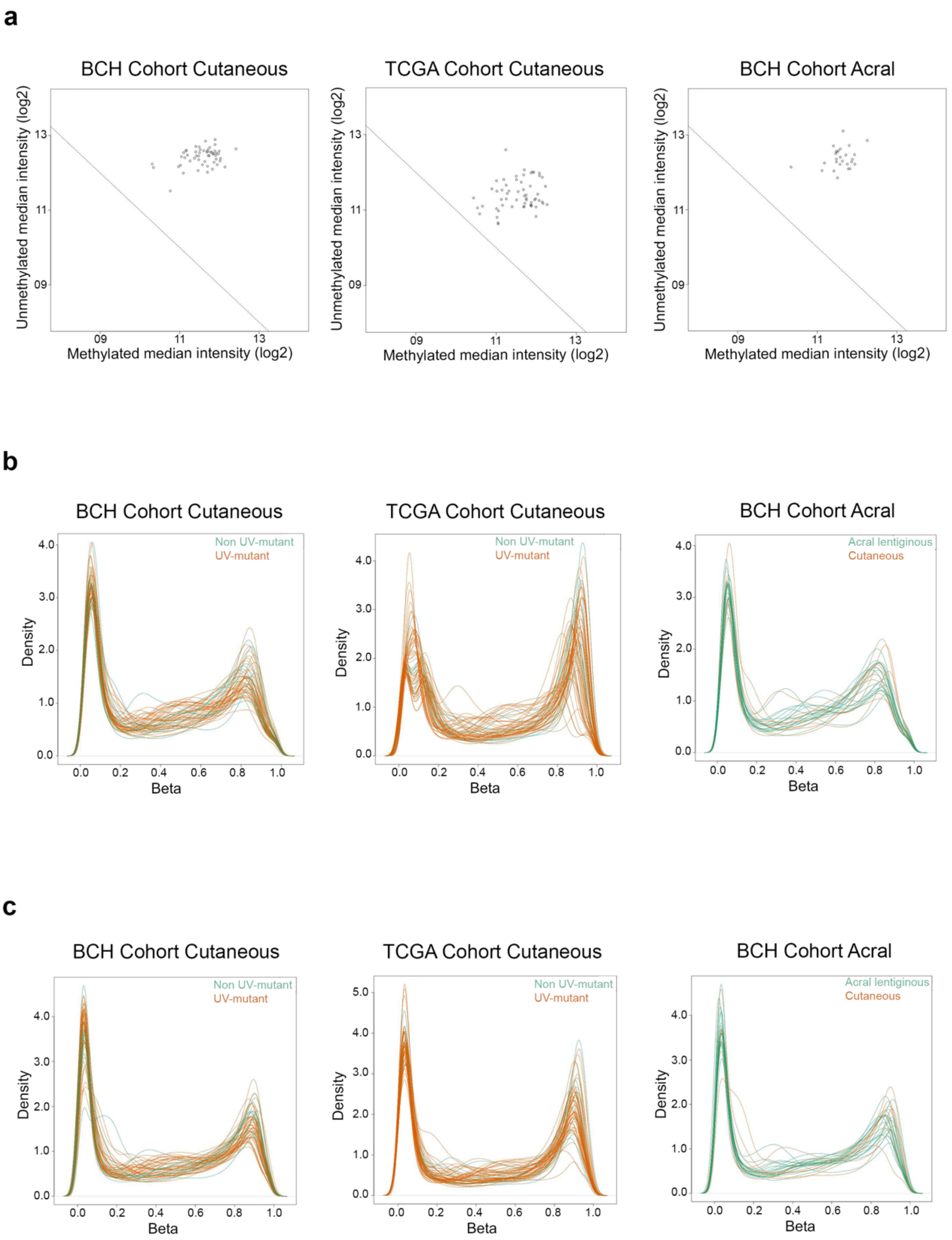


**Supplementary Figure 4**: Quality control of 450K methylation data. a) The plots show that all samples passed quality control (above diagonal threshold line) in each of the three indicated cohorts (BCH Cohort Cutaneous= 54, TCGA Cohort Cutaneous= 58 and BCH Cohort Acral= 21) ; b) Density plots of the beta methylation values for BCH-cutaneous, TCGA-cutaneous and BCH-acral datasets. In BCH-cutaneous and TCGA-cutaneous plots, orange and green densities represent samples harboring or not the UV signature, respectively. In BCH-acral, orange and green densities represent cutaneous (not harboring UV signature) and acral melanomas, respectively. c) Density plots of (b) after FunNorm normalization.

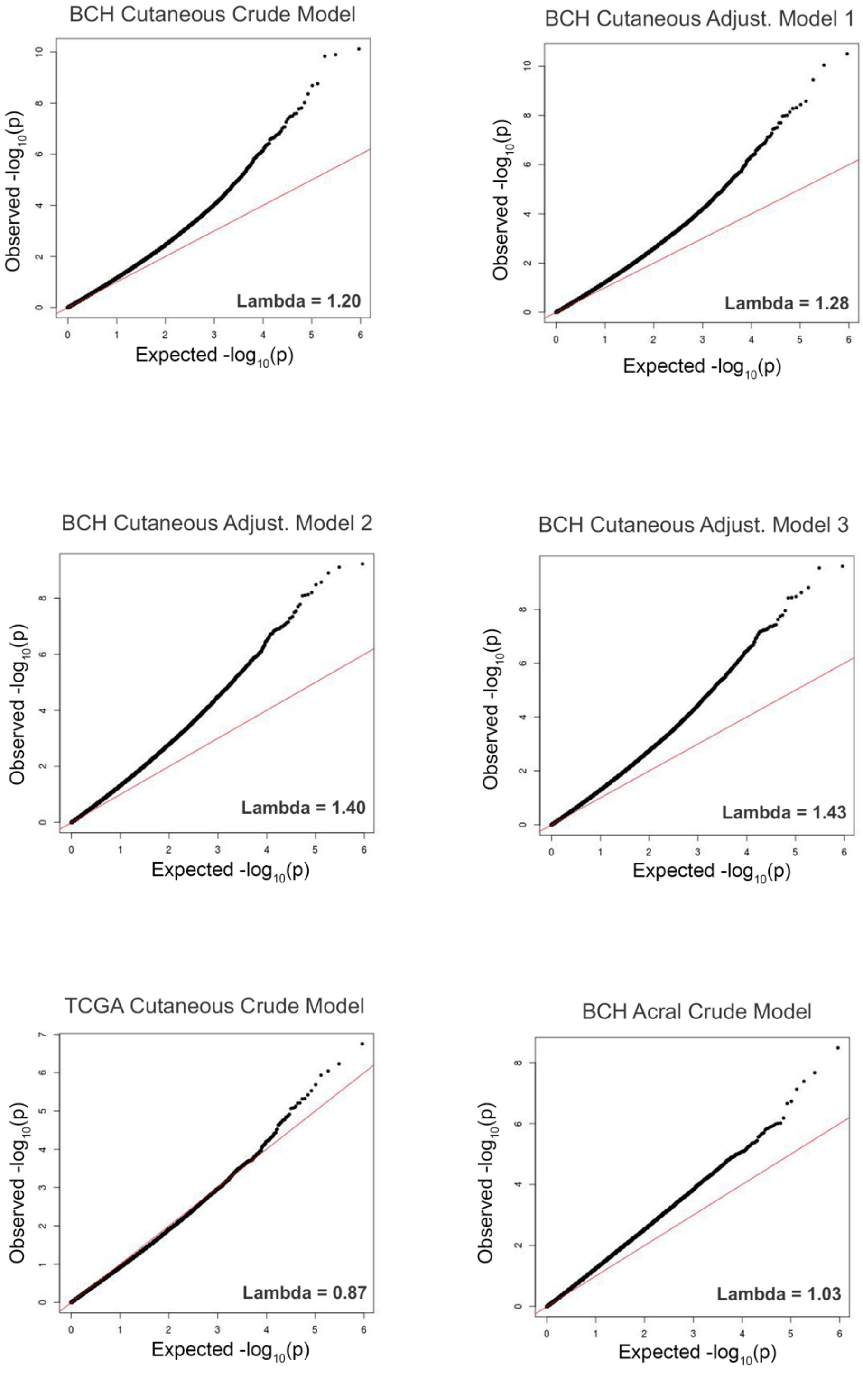


**Supplementary Figure 5**: Q-Q plots and lambda genomic inflation values of the statistical models used in BCH-cutaneous (n= 54), TCGA-cutaneous (n=58) and BCH-acral melanoma (n=17) cohorts. Crude and adjusted models are described in Methods.

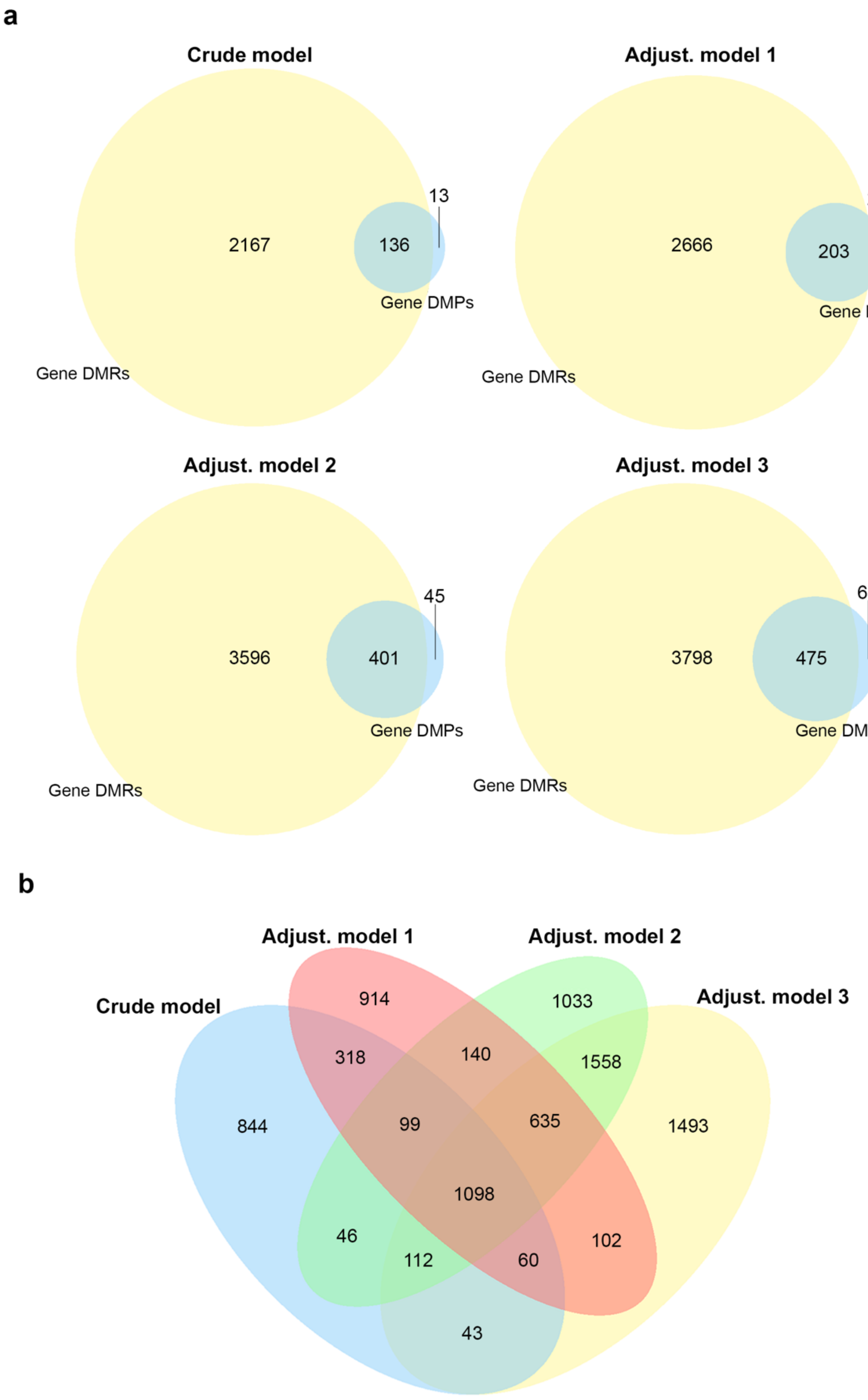


**Supplementary Figure 6**: Comparison of results yielded from the tested crude and adjusted statistical models as well as the two approaches used for the analyses, DMP and DMR. a) Approximately 90% of DMP-based genes overlapped with DMR-based genes across all tested models. b) Venn diagram showing DMR overlaps among the 4 statistical models.

**Supplementary References**


1. Bertrand, D. *et al.* ConsensusDriver Improves upon Individual Algorithms for Predicting Driver Alterations in Different Cancer Types and Individual Patients. *Cancer Res* **78**, 290-301 (2018).
2. Trucco, L.D. *et al.* Ultraviolet radiation-induced DNA damage is prognostic for outcome in melanoma. *Nat Med* **25**, 221-224 (2019).

**Description of Additional Supplementary Files**

Cutaneous and acral melanoma cross-OMICs reveals prognostic cancer drivers associated with pathobiology and ultraviolet exposure
Vicente A.L.S.A. *et al*.

**Supplementary Data 1**. Systematic literature search conducted in the PubMed database until May 2021 in order to find out studies that investigated the DNA methylome profile of melanoma patients.

**Supplementary Data 2**. DMRs in relation to UV exposure in BCH-cutaneous crude model.

**Supplementary Data 3**. DMRs in relation to UV exposure in BCH-cutaneous adjusted for sex.

**Supplementary Data 4**. DMRs in relation to UV exposure in BCH-cutaneous adjusted for sex and age at diagnosis.

**Supplementary Data 5**. DMRs in relation to UV exposure in BCH-cutaneous adjusted for sex, age at diagnosis and tumor type (primary or metastatic).

**Supplementary Data 6**. DMRs in relation to UV exposure in TCGA-cutaneous crude model.

**Supplementary Data 7**. All CpGs differentially methylated comparing non UV-mutant *versus* UV-mutant in crude model that passed the filtration steps described in Supplementary Fig. 1a and used in pathway and heatmap cluster analysis (Figure 3a and 3c) in BCH.

**Supplementary Data 8**. All CpGs differentially methylated comparing non UV-mutant *versus* UV-mutant in crude model that passed the filtration steps described in Supplementary Fig. 1a and used in pathway and heatmap cluster analysis (Figure 3a and 3c) in TCGA.

**Supplementary Data 9**. Jensen Ontology analysis using genes prioritized in Supplementary Fig. 1a and described in Supplementary Data 7 in BCH cohort using Enrich-r website. P-value was delivered from two-sided Fisher's exact test.

**Supplementary Data 10**. Jensen Ontology analysis using genes prioritized in Supplementary Fig. 1a and described in Supplementary Data 8 in TCGA cohort using Enrich-r website. P-value was delivered from two-sided Fisher's exact test.

**Supplementary Data 11**. KEGG pathway analysis using genes prioritized in Supplementary Fig. 1a and described in Supplementary Data 7 in BCH cohort using Enrich-r website. P-value was delivered from two-sided Fisher's exact test.

**Supplementary Data 12**. KEGG pathway analysis using genes prioritized in Supplementary Fig. 1a and described in Supplementary Data 8 in TCGA cohort using Enrich-r website. P-value was delivered from two-sided Fisher's exact test.

**Supplementary Data 13**. KEGG Pathway analysis using CpGs prioritized in Supplementary Fig. 1a and described in Supplementary Data 7 in BCH cohort using missMethyl package, which adjusts for the number of CpG associated with each gene. P-value was delivered from two-sided Fisher's exact test.

**Supplementary Data 14**. KEGG Pathway analysis using CpGs prioritized in Supplementary Fig. 1a and described in Supplementary Data 8 in TCGA cohort using missMethyl package, which adjusts for the number of CpG associated with each gene. P-value was delivered from two-sided Fisher's exact test.

**Supplementary Data 15**. Differentially expressed genes comparing non UV-mutant and UV-mutant in TCGA cohort.

**Supplementary Data 16**. Jensen Ontology analysis using differentially expressed genes described in Supplementary Data 15 in TCGA cohort using Enrich-r website. P-value was delivered from two-sided Fisher's exact test.

**Supplementary Data 17**. KEGG Pathway analysis using differentially expressed genes described in Supplementary Data 15 in TCGA cohort using Enrich-r website. P-value was delivered from two-sided Fisher's exact test.

**Supplementary Data 18**. 458 CpGs in common between BCH and TCGA.

**Supplementary Data 19**. Meta-analysis of DMRs across the BCH and TCGA datasets considering FDR-adjusted $p < 0.05$ and DMRs with at least 3 CpGs. The approach used is fixed effects inverse variance-weighted meta-analysis. P-value was derived from two-sided test based on Z-score (obtained from the direction of effect and P-value observed in each DMR) and the standard normal cumulative distribution function.

**Supplementary Data 20**. Meta-analysis of DMRs across the BCH and TCGA datasets considering Bonferroni-adjusted $p < 0.05$ and DMRs with at least 3 CpGs. The approach used is fixed effects inverse variance-weighted meta-analysis. P-value was derived from two-sided test based on Z-score (obtained from the direction of effect and P-value observed in each DMR) and the standard normal cumulative distribution function.

**Supplementary Data 21**. Association between DNA methylation and melanoma-specific survival in BCH and TCGA cohorts of CpGs common between the two cohorts, after the filters applied in Supplementary Fig. 1b. P-values were derived from log rank test.

**Supplementary Data 22**. Primers used for pyrosequencing validation of the *TAPBP* gene. "For", "Rev" and "Seq" denote forward, reverse and sequencing primers, respectively.

**Supplementary Data 23.** Twenty-five most informative CpGs and transcripts using LASSO penalization for integrative analysis in the TCGA cohort.

**Supplementary Data 24.** DMRs in BCH cohort comparing acral *versus* cutaneous in non UV-exposed melanoma patients.

**Supplementary Data 25.** All CpGs differentially methylated comparing acral *versus* cutaneous BCH melanoma patients that passed the filtration steps described in Supplementary Fig. 2e and used in gene ontology analysis (Figure 6d).

**Supplementary Data 26**. Jensen Ontology analysis using genes prioritized in Supplementary Fig. 2e and described in Supplementary Data 25 in BCH-acral cohort using Enrich-r website. P-value was delivered from two-sided Fisher's exact test.

**Supplementary Data 27**. KEGG pathway analysis using genes prioritized in Supplementary Fig. 2e and described in Supplementary Data 25 in BCH-acral using Enrich-r website. P-value was delivered from two-sided Fisher's exact test.

**Supplementary Data 28**. KEGG Pathway analysis using CpGs prioritized in Supplementary Fig. 2e and described in Supplementary Data 25 in BCH cohort using missMethyl package, which adjusts for the number of CpG associated with each gene. P-value was delivered from two-sided Fisher's exact test.

**Supplementary Data 29**. Association between DNA methylation and melanoma-specific survival of genes frequently mutated in response to UV exposure[1]. P-values were derived from log rank test. Adjustment for multiple testing was done using FDR.

### Supplementary Reference

1- Trucco, L.D. *et al.* Ultraviolet radiation-induced DNA damage is prognostic for outcome in melanoma. *Nat Med* **25**, 221-224 (2019).
